# Evaluating the impact of a gambling harm focused educational intervention on medical students’ self-reported confidence: a non-randomised, single-group pre-post study

**DOI:** 10.64898/2026.09.01.26361237

**Authors:** Marriam Albukai, Andrew Lovell, Ben Jones, Kishan Patel

**Author notes:** **Correspondence to**: Marriam Albukai.

## Abstract

**Background:** Gambling harm is a significant public health concern that is systematically under-recognised in clinical practice. Despite the recent inclusion of gambling disorder in the General Medical Council’s Medical Licensing Assessment content map, gambling harm has been largely absent from undergraduate medical education in the United Kingdom, and structured evaluations of gambling harm teaching delivered to medical students have not, to our knowledge, been reported.

**Methods:** A single-group, pre-post study of a teaching session on gambling harm was conducted across three sequential cohorts of Year 4 medical students at King’s College London during the 2025-2026 academic year. Outcome measures were collected immediately after the teaching with no follow-up. The session was delivered online by Gambling Harm UK, a registered UK charity, and comprised lived experience testimony and teaching with public health and clinical components. Self-reported confidence across six domains was assessed pre-and post-session on a five-point scale, alongside nine post-session attitudinal statements. Paired confidence data were analysed using the Wilcoxon signed-rank test with Hodges-Lehmann estimates of the median paired difference, with a Bonferroni-corrected significance threshold of p < 0.008 applied within each cohort.

**Results:** Sixty-eight paired responses were analysed (completion rates of 28%, 24%, and 32%). Statistically significant improvements in self-reported confidence were observed across all six domains in each cohort. Median confidence rose from’slightly confident’ before the session to’quite confident’ afterwards, with median paired improvements of 1.5 to 2.5 points (all p ≤ 0.001). Post-session attitudinal responses were positive across all nine items, with lived experience the most strongly endorsed.

**Conclusions:** A single online teaching session on gambling harm was associated with significant and consistent improvements in self-reported confidence in recognising and responding to gambling harm across three cohorts of medical students. Whether these gains translate into changes in clinical behaviour requires longer-term, multi-site evaluation with behavioural outcome measures. Medical schools should consider how gambling harm teaching might fit within their curricula and evaluate its introduction, with the aim of normalising asking about gambling in routine social history taking alongside alcohol and smoking.

## Background

Gambling harm is a significant and growing public health concern. Recent estimates suggest that around one in seven adults who gamble experience some degree of harm themselves^1^, and this harm is not evenly distributed, with socio-economically deprived and ethnic minority communities affected disproportionately^2–4^. Individuals experiencing gambling harm are at substantially higher risk of suicide^5–7^. Harm also extends beyond the individual: for each person experiencing gambling problems, an average of six others are affected^8,9^, with partners and children experiencing financial harm and family violence^10–12^. Population-level evidence suggests that most total gambling harm, for at least some types of harm, may be attributable to the larger group of individuals at low-to-moderate risk rather than the smaller number meeting threshold criteria for gambling disorder^13–15^. This underscores the need for clinical approaches that recognise gambling harm across the full spectrum of severity, and for teaching that equips future clinicians to identify harms and refer patients to relevant services.

Despite the severity of its harms, gambling is systematically under-recognised in clinical practice. This is driven partially by the normalisation of gambling including through industry-led advertising and the integration of betting into sport^16,17^, and by stigma, which deters people from seeking help or disclosing their gambling^18,19^. Individuals experiencing gambling harm frequently present to healthcare services with its consequences rather than the gambling itself, across settings ranging from general practice to mental health services to emergency care^20–22^. However, the gambling underlying these presentations usually goes undetected. Unlike alcohol, smoking, and drug use, it is rarely included in routine social history taking, and patients seldom raise it themselves^21,22^. Consistent with this, gambling is infrequently recorded in routine clinical data, even in primary care where most coding occurs^23^. Furthermore, when people do seek help for the gambling itself, they often reach services once harm has become severe^21,23^, which represents a significant missed opportunity for early identification and intervention^24^.

Gambling harm has recently gained more recognition within medical education in the United Kingdom. Blythe and van Schalkwyk advocated for the integration of gambling harms into medical school curricula^24^. The recent revision of the content of the Medical Licensing Assessment (MLA) established gambling disorder as part of the core clinical knowledge that newly qualified UK doctors must hold to practise safely, effective from September 2026^25^. In a conference abstract, Rice and Mansi reported an online teaching session on gambling harm developed in collaboration with GamCare and delivered to an earlier cohort of Year 4 medical students at King’s College London, though no structured learning objectives or formal evaluation were reported^26^. To our knowledge, no structured evaluation of gambling harm teaching for medical students has been published.

The present study evaluates a teaching session on gambling harm delivered to a cohort of Year 4 medical students at King’s College London by Gambling Harm UK, a registered UK charity. The session was designed and delivered by individuals with lived experience of gambling harm and expertise in public health, and drew on a public health framing and on contact-based pedagogy, which has an established evidence base in stigma reduction^27,28^. We sought to assess whether the session improved self-reported confidence in recognising and responding to gambling harm, and to characterise post-session attitudes toward gambling harm as a clinical and public health issue. More broadly, the session aimed to support the normalisation of asking about gambling in routine social history taking, alongside alcohol and smoking.

## Methods

### Study design and reporting

This was a single-group, pre-post evaluation conducted across three sequential cohorts. Randomisation, stratification, or blinding to the intervention or the evaluative purpose of the surveys was not applicable with this design. The study was reported in accordance with the TREND statement for non-randomised evaluations of behavioural and public health interventions^29^. The completed checklist is provided in Supplementary File 1.

### Participants

Invited participants were Year 4 medical students on the MBBS programme at King’s College London who were undertaking their psychiatry rotation. No other eligibility criteria were used. Attendance was timetabled but not formally enforced. All attendees received the same intervention. Vevox participants were defined as attendees who joined the platform Vevox by scanning a QR code displayed at the start of each session, regardless of whether they completed any survey items. Completion rates were defined as the proportion of Vevox participants who completed both pre-and post-teaching surveys. No a priori power calculation was performed.

### Objectives

The primary objective was to evaluate whether a single educational session on gambling harm improved medical students’ self-reported confidence in recognising and responding to gambling harm. The secondary objective was to characterise students’ post-session attitudes toward the relevance of gambling harm to clinical practice and their perceptions of the session quality and format. Qualitative feedback was collected as an exploratory aim.

### Intervention

A single online session of approximately one hour and forty-five minutes was delivered to each of three cohorts across the 2025-2026 academic year. Each session comprised the pre-teaching survey, the teaching, and the post-teaching survey.

The session was delivered live online via Microsoft Teams to each cohort as a single group by two representatives of Gambling Harm UK. Both have lived experience of gambling harm, one as an affected other, and one as an individual with lived experience of gambling disorder. The former is also a public health doctor.

The session combined personal accounts of gambling harm and its impact on individuals and affected others with structured teaching and interactive audience polling via Vevox. Teaching content covered the epidemiology of gambling harm, population-level impacts and health economics, the public health approach to gambling including advertising and product design, stigma and language, inequalities, and dopamine-agonist induced gambling. Clinical components addressed how gambling harm presents in practice, why it is often missed, how to ask about gambling, and how to refer patients to appropriate services.

The session content was iteratively refined between cohorts while retaining the same core structure. Cohort 2 included additional video content on the impact of gambling on children and families. Cohort 3 included three case-based discussions.

### Outcome measures and data collection

No validated instrument for measuring confidence in recognising or responding to gambling harm was identified in the literature, so the survey items were developed specifically for this study and informed by the session’s learning objectives. The six confidence items were rated on a five-point scale (1 = not at all confident, 5 = extremely confident), and the nine attitudinal statements on a five-point agreement scale (1 = strongly disagree, 5 = strongly agree). For analysis and presentation, the attitudinal statements were grouped post hoc into four themes: perceived importance of gambling harm, session quality and format, lived experience, and intention and endorsement of the session. Two open-ended questions captured qualitative feedback on the most valuable aspects of the session and suggestions for improvement. The full instrument is provided in Supplementary File 2.

The instrument was administered as a single continuous Vevox survey within each session to enable anonymous matching of paired responses. Responses were only captured when a participant submitted the survey.

Before the teaching began, students completed six consent statements and the six pre-session confidence items. After the teaching concluded, they completed the six post-session confidence items, nine attitudinal statements, and two optional open-ended questions. Participation was voluntary, and students were informed that responses would be used for evaluation, that all data were anonymous, and that declining to take part carried no negative consequences. No reimbursements or incentives were provided.

## Data analysis

The unit of analysis was the individual student. Cohorts were analysed separately to assess the consistency of findings. Analysis was conducted by an author who had no role in intervention design or delivery.

Pre-and post-session confidence ratings were summarised using medians and interquartile ranges. The Wilcoxon signed-rank test was used as the primary analysis to compare pre-and post-session ratings for each paired confidence question. The magnitude of change in confidence was quantified using the Hodges-Lehmann estimate of the median paired difference with its 95% confidence interval^30^. This estimator assumes that differences between adjacent scale points are comparable. Participants with missing responses for a given question were excluded from that question’s analysis, so paired sample sizes could vary across questions. A Bonferroni-corrected statistical significance threshold of p < 0.008 was applied to account for multiple testing across the six confidence questions within each cohort.

Post-session attitudinal responses were summarised using medians, interquartile ranges, and the proportion of respondents selecting ‘Agree’ or ‘Strongly agree’. Participants with missing responses for a given statement were excluded from that statement’s summary.

Qualitative responses were reviewed and organised into recurring themes by cohort. This process was descriptive and intended to complement the quantitative findings rather than constitute a formal qualitative analysis.

Analyses were conducted using R (version 4.4.2)^31^ with the readxl, dplyr, and ggplot2^32^ packages.

## Ethics statement

The Education Ethics Review Process of Imperial College London gave ethical approval for this work (reference EERP2425-142, 2 June 2025), where the evaluation protocol was developed. The Research Governance, Ethics and Integrity department of King’s College London gave ethical approval for this work at that site, where the sessions were delivered (20 October 2025). All participants provided informed consent via the Vevox platform prior to completing the pre-session survey.

## Results

### Participant flow

Three sessions were delivered on 31 October 2025 (Cohort 1), 23 January 2026 (Cohort 2), and 27 March 2026 (Cohort 3), timetabled for the full psychiatry rotation cohort. Exact attendance was not recorded. Participation in the in-session Vevox polling was 85, 83, and 74 students in Cohorts 1, 2, and 3 respectively, of whom 24, 20, and 24 completed both the pre-and post-session surveys, giving completion rates of 28%, 24%, and 32%. Across all three cohorts, 68 of 242 Vevox participants completed both surveys, an overall completion rate of 28%. These 68 paired respondents formed the analysed sample. Figure 1 summarises participant flow.

**Figure 1.**
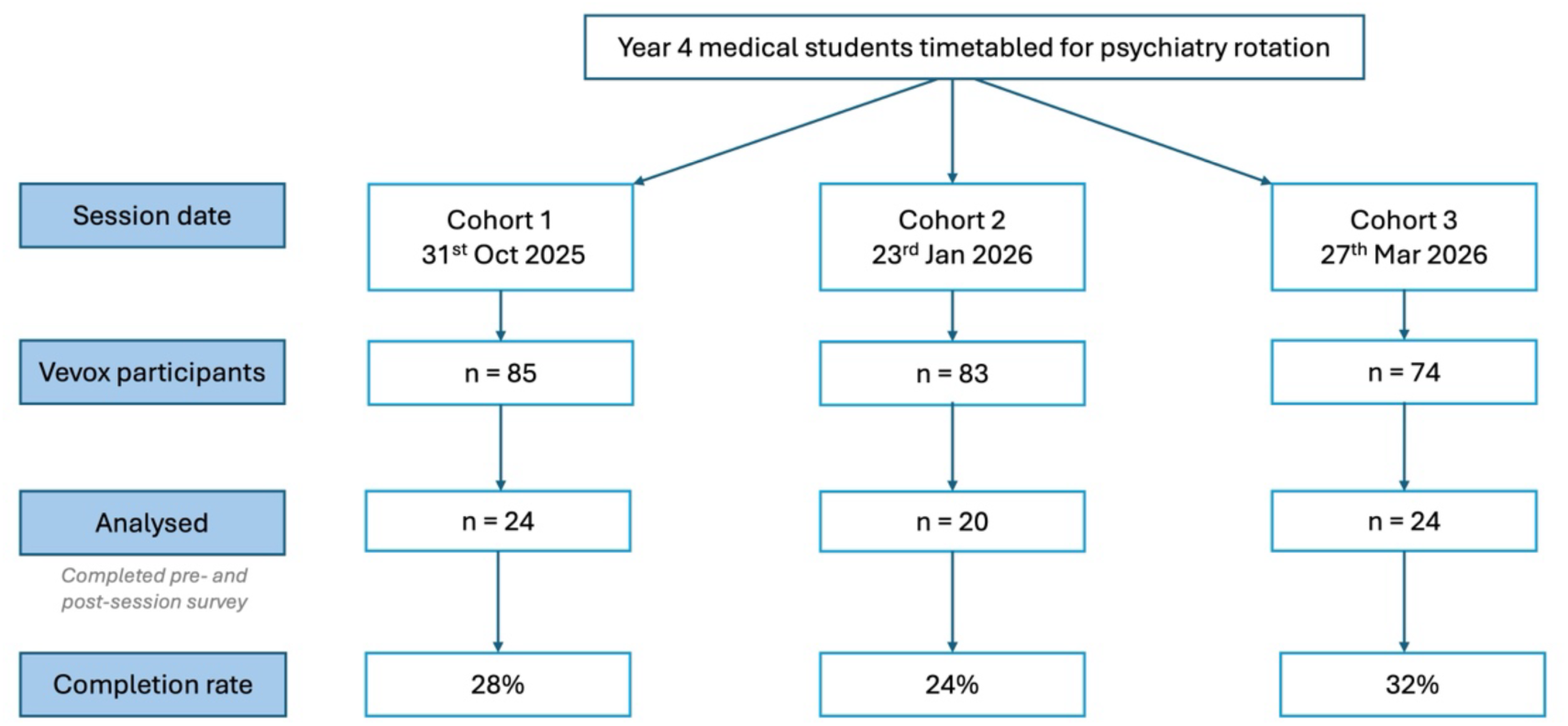
Participant flow through each stage of the study. All attendees received the same teaching session with no comparison group. Outcome measures were collected immediately after the teaching with no follow-up period. Completion rate is the proportion of Vevox participants who submitted both the preand post-teaching surveys.

No unplanned deviations from the described methods occurred in any session. Outcome measures were collected immediately after the teaching without a longitudinal follow-up period. No demographic data were collected, so pre-session confidence ratings were the only available baseline characterisation.

### Pre-and post-session self-reported confidence

Paired sample sizes for the six confidence items ranged from 23 to 24 in Cohort 1 and 19 to 20 in Cohort 2 and were complete (24) in Cohort 3.

Prior to the teaching, participants in all three cohorts reported low confidence across all six domains, with responses concentrated in the ‘Not at all confident’ and ‘Slightly confident’ categories (Figure 2). All pre-session medians were at or below ‘Slightly confident’ (Table 1). Following the teaching, median confidence was ‘Quite confident’ across all six domains in all three cohorts.

**Figure 2.**
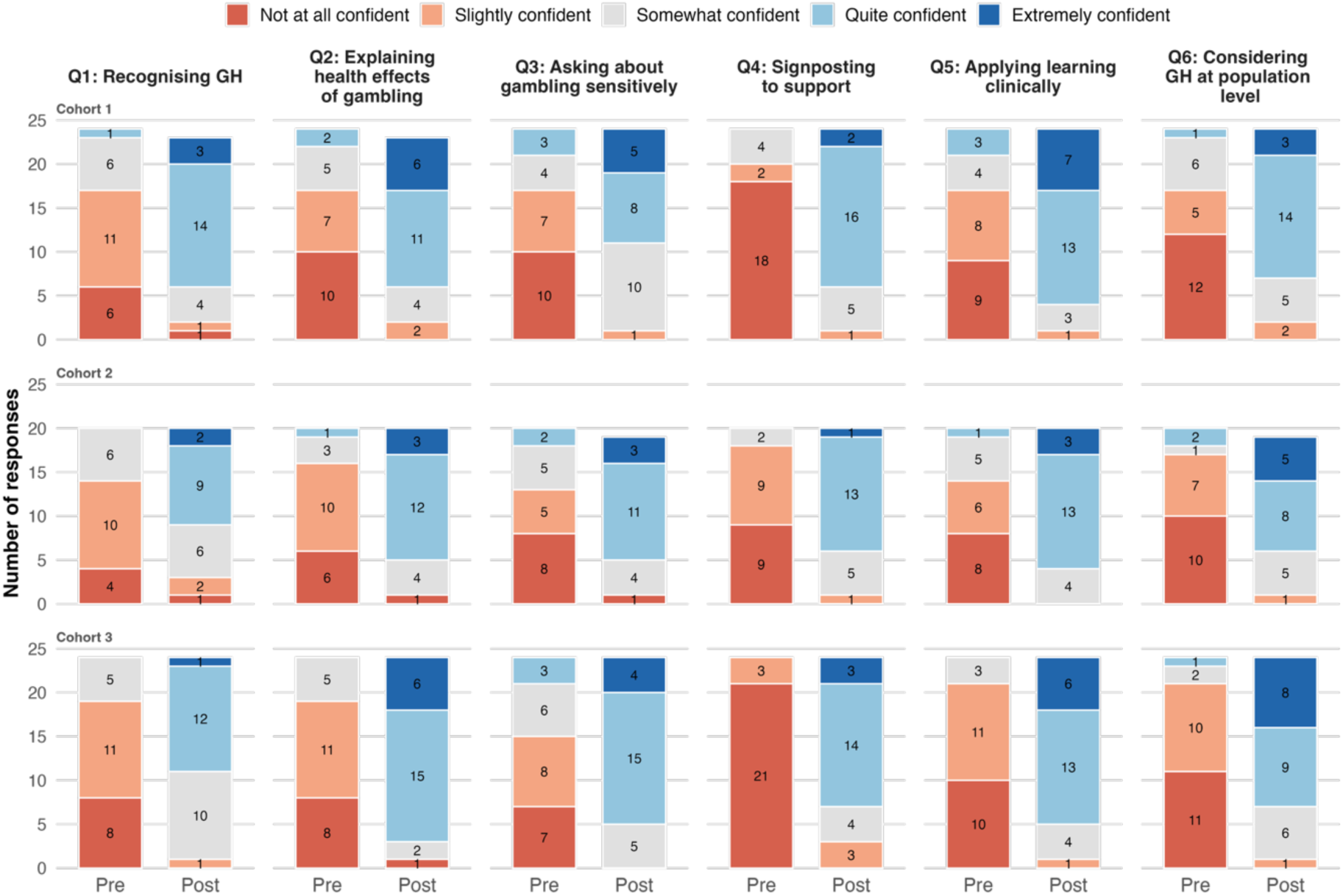
Distribution of pre-and post-session responses to the six confidence questions, by cohort. Responses were rated on a five-point scale (1 = not at all confident, 2 = slightly confident, 3 = somewhat confident, 4 = quite confident, 5 = extremely confident); bars diverge from the midpoint of the “somewhat confident” category. Numbers within bars are respondent counts. All pre–post comparisons were significant (Wilcoxon signed-rank test, all p ≤ 0.001). GH, gambling harm.

**Table 1.** Pre-and post-session self-reported confidence in addressing gambling harm across three cohorts.

| Question | Cohort | n | Pre-session<br>Median<br>(IQR) | Post-session<br>Median<br>(IQR) | Median<br>difference<br>(95% CI) | p |
| --- | --- | --- | --- | --- | --- | --- |
| Recognising gambling harm in patients | Cohort 1 | 23 | 2.0 (2.0–3.0) | 4.0 (3.5–4.0) | 2.0 (1.5–2.0) | <0.001 |
|  | Cohort 2 | 20 | 2.0 (2.0–3.0) | 4.0 (3.0–4.0) | 1.5 (1.0–2.0) | 0.001 |
|  | Cohort 3 | 24 | 2.0 (1.0–2.0) | 4.0 (3.0–4.0) | 1.5 (1.5–2.0) | <0.001 |
| Explaining health effects of gambling harm | Cohort 1 | 23 | 2.0 (1.0–3.0) | 4.0 (3.5–4.5) | 2.0 (2.0–2.5) | <0.001 |
|  | Cohort 2 | 20 | 2.0 (1.0–2.0) | 4.0 (3.8–4.0) | 2.0 (1.5–2.5) | <0.001 |
|  | Cohort 3 | 24 | 2.0 (1.0–2.0) | 4.0 (4.0–4.2) | 2.5 (2.0–2.5) | <0.001 |
| Asking about gambling sensitively and non-judgementally | Cohort 1 | 24 | 2.0 (1.0–3.0) | 4.0 (3.0–4.0) | 2.0 (1.5–2.0) | <0.001 |
|  | Cohort 2 | 19 | 2.0 (1.0–3.0) | 4.0 (3.5–4.0) | 2.0 (1.5–2.5) | <0.001 |
|  | Cohort 3 | 24 | 2.0 (1.0–3.0) | 4.0 (4.0–4.0) | 2.0 (1.5–2.5) | <0.001 |
| Identifying referral or support options | Cohort 1 | 24 | 1.0 (1.0–1.2) | 4.0 (3.8–4.0) | 2.5 (2.0–3.0) | <0.001 |
|  | Cohort 2 | 20 | 2.0 (1.0–2.0) | 4.0 (3.0–4.0) | 2.0 (2.0–2.5) | <0.001 |
|  | Cohort 3 | 24 | 1.0 (1.0–1.0) | 4.0 (3.0–4.0) | 2.5 (2.5–3.0) | <0.001 |
| Applying learning in clinical interactions | Cohort 1 | 24 | 2.0 (1.0–3.0) | 4.0 (4.0–5.0) | 2.0 (2.0–2.5) | <0.001 |
|  | Cohort 2 | 20 | 2.0 (1.0–3.0) | 4.0 (4.0–4.0) | 2.0 (1.5–2.5) | <0.001 |
|  | Cohort 3 | 24 | 2.0 (1.0–2.0) | 4.0 (4.0–4.2) | 2.5 (2.0–3.0) | <0.001 |
| Considering gambling harm in population-level discussions | Cohort 1 | 24 | 1.5 (1.0–3.0) | 4.0 (3.0–4.0) | 2.0 (2.0–2.5) | <0.001 |
|  | Cohort 2 | 19 | 1.0 (1.0–2.0) | 4.0 (3.0–4.5) | 2.5 (1.5–3.0) | <0.001 |
|  | Cohort 3 | 24 | 2.0 (1.0–2.0) | 4.0 (3.0–5.0) | 2.5 (2.0–3.0) | <0.001 |

Responses were rated on a five-point Likert scale: 1 = Not at all confident, 2 = Slightly confident, 3 = Somewhat confident, 4 = Quite confident, 5 = Extremely confident.
Median difference was the Hodges-Lehmann estimate of the paired difference (post minus pre), with its 95% confidence interval.
Quantiles were computed by linear interpolation and may take non-integer values.
Pre- and post-session differences within each cohort were assessed using the Wilcoxon signed-rank test.
Participants with missing responses for an item were excluded from that item's analysis.
With six items tested per cohort, the Bonferroni-corrected significance threshold was $p < 0.008$ .
n = number of paired responses; IQR = interquartile range; CI = confidence interval.

The Wilcoxon signed-rank test indicated statistically significant increases in self-reported confidence across all six domains in all three cohorts (all p ≤ 0.001, all below the Bonferroni-corrected threshold of p < 0.008). Median increases ranged from 1.5 to 2.5 points across domains and cohorts.

Five of the 68 paired respondents (7%) recorded lower post-session confidence on at least one item: one in Cohort 1, four in Cohort 2, and none in Cohort 3. These accounted for seven item-level decreases, six of a single scale point and one of two points.

### Post-session attitudinal responses

Post-session attitudinal responses were consistently positive across all three cohorts, with ‘Strongly agree’ the most common response to almost every item, followed by ‘Agree’ (Figure 3). Across the perceived importance items, participants endorsed that gambling harm is a modifiable risk factor and a neglected public health concern, and that healthcare professionals have a role in recognising and responding to it. The value of lived experience in enhancing understanding received the highest number of ‘Strongly agree’ responses of any single item. Participants consistently indicated their intention to apply learning and would recommend the session to peers. Responses outside the ‘Agree’ and ‘Strongly agree’ categories were uncommon and concentrated on session length, for which ‘Agree’ rather than ‘Strongly agree’ predominated, four participants selected ‘Neither’ agree nor disagree, and two disagreed. Isolated ‘Neither’ agree nor disagree responses were also recorded for the modifiable risk factor, materials, and interaction items, and one participant disagreed that lived experience enhanced their understanding. Medians, interquartile ranges, and proportions agreeing are provided in Supplementary Table 1.

**Figure 2.**
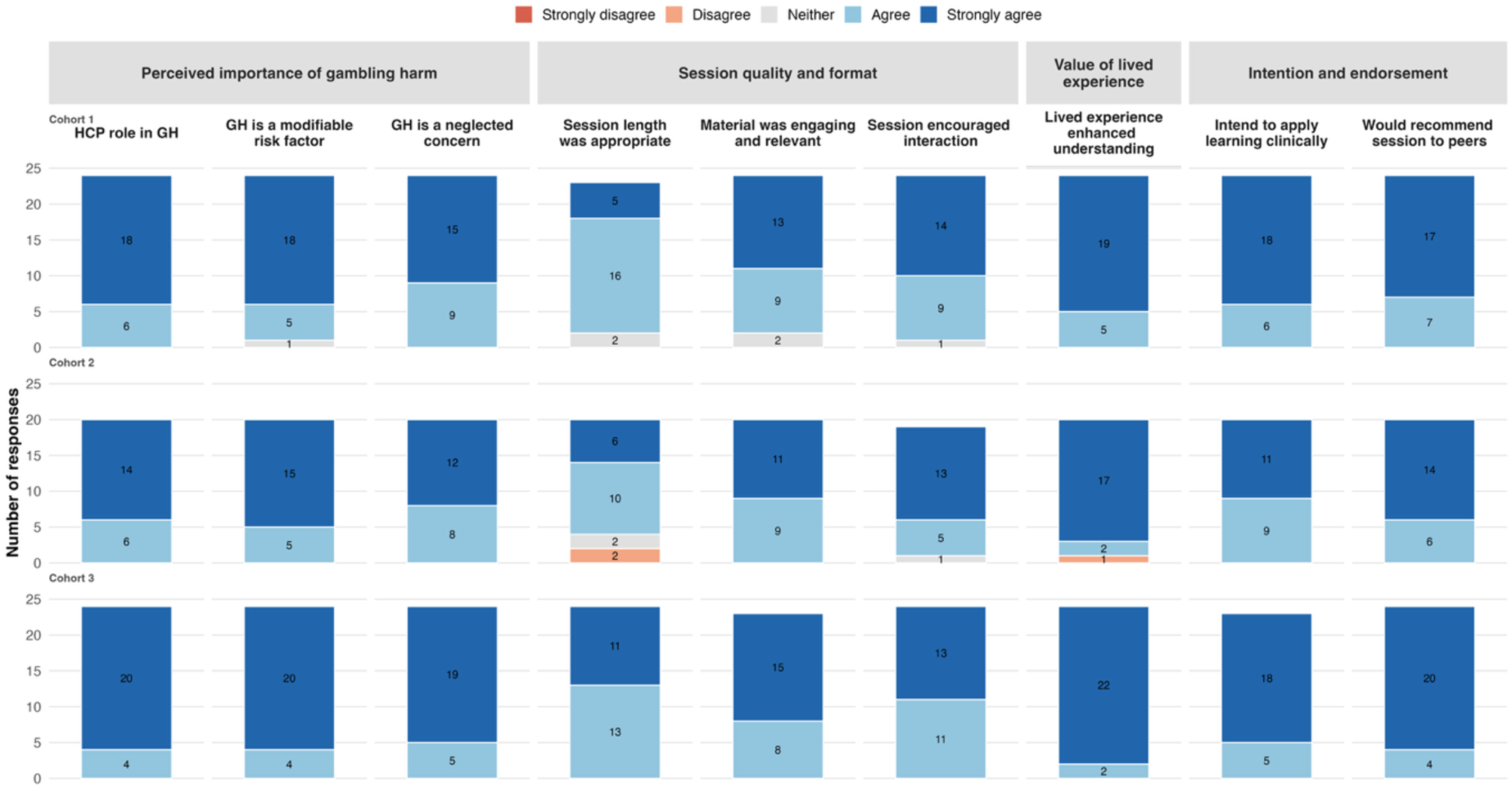
Distribution of post-session attitudinal responses across the three cohorts, grouped by theme: perceived importance of gambling harm (statements 1-3), session quality and format (statements 4-6), value of lived experience (statement 7), and intention and endorsement of the session (statements 8-9). Numbers within bars indicate respondent counts. Response categories ranged from Strongly disagree to Strongly agree. HCP= Health care professionals. GH = gambling harm.

### Qualitative responses

Fifty-five respondents commented on what they found most valuable (17, 17, and 21 from Cohorts 1, 2, and 3 respectively) and 33 offered suggestions for improvement (8, 10, and 15 across the three cohorts respectively). Most valuable aspects

Lived experience accounts were identified as the most valuable element of the session by most respondents in every cohort, described as “engaging”, “memorable”, and “impactful”, and “way more eye opening than facts on a lecture slide”. Several valued hearing from both someone who had gambled and a family member affected by another’s gambling. One respondent offered a counterpoint, stating that they “didn’t gain much from the personal stories personally”, indicating that the format did not resonate uniformly across all students.

Beyond the personal accounts, the teaching was described as “interactive” and “insightful”, and by one respondent as “one of the best teachings we’ve had”. Respondents across all cohorts valued learning how to recognise gambling harm and how to ask about it in consultation using non-stigmatising language, with one noting that the session was “the first time we were taught about all the health / social implications of gambling”. Several specifically welcomed the case-based discussions as an opportunity to apply their new knowledge.

The most consistent suggestion, raised in all three cohorts, was for greater opportunity to practise clinical skills. Respondents proposed role play, case vignettes, simulated consultations, and worked examples of “how a patient may present”, and asked for more interactivity and small group discussion, with several expressing a preference for in-person delivery. Comments on pacing also recurred across cohorts, including condensing the session, introducing a longer mid-session break, and reducing the time spent on population-level statistics. Others requested more detail on interventions and care pathways for patients, a summary sheet to take away, and resolution of technical issues affecting video playback and survey functionality.

No adverse events or indications of distress were reported during or following any session.

## Discussion

This study evaluated a teaching session on gambling harm, delivered by individuals with lived experience and public health expertise, among Year 4 medical students at King’s College London. The primary hypothesis of improved post-session self-reported confidence across all six domains was supported in all three cohorts. Median confidence rose from ‘slightly confident’ before the session to ‘quite confident’ afterwards, across all six domains among the 68 students (28% of Vevox participants) who completed both surveys. Post-session attitudinal responses were strongly positive, and qualitative feedback consistently highlighted the value of lived experience testimony.

### Interpretation of findings

Baseline confidence was low across all domains and cohorts, with identifying referral or support options and considering gambling harm in population-level discussions consistently among the weakest areas. This uniformly low baseline suggests that students at this institution had received little or no prior teaching on gambling harm as a clinical topic. The qualitative data echo this, with one respondent describing the session as “the first time” they had been taught about the harms of gambling, consistent with the limited attention gambling has received in undergraduate curricula^24^ despite its recent inclusion in the MLA content map^25^. The low baseline confidence also means participants had substantial room to improve, which may have contributed to the size of the observed gains.

Two features of the session may explain its effects. First, the lived experience testimony appears to have created the intended affective connection by making gambling harm feel real rather than abstract. The qualitative data are consistent with this mechanism, as lived experience was identified as the most valuable element in every cohort. Second, the structured teaching gave students a practical framework for recognising how gambling harm presents, asking about it sensitively, and knowing where to refer, set within the public health context that explains why these harms are common and often missed. The combination of affective engagement and structured content may explain the breadth of the effect across all six domains.

### Comparison with existing evidence

To our knowledge, this is the first structured evaluation of gambling harm teaching delivered to medical students. The most closely comparable work is the session described by Rice and Mansi, which was also delivered online to Year 4 students at the same institution. It reached an earlier cohort, was developed in collaboration with a different organisation (GamCare), and was delivered by different individuals. Their informal observations are consistent with the present findings, but no structured learning objectives or formal evaluation were reported^26^. Thomas et al. found that lived experience performances on gambling harm increased confidence to explore gambling among allied professionals and community members in Australia, although their study used a post-only design and a different target population^33^. More broadly, the magnitude of confidence gain observed here is consistent with pre-post evaluations of teaching on other clinical topics that are stigmatised and absent from routine history taking, such as intimate partner violence and child safeguarding^34^.

### Implementation, barriers and future iterations

The consistency of confidence shifts and qualitative themes across cohorts provides some evidence of reproducibility, although the design cannot attribute the effect to specific components. Participant feedback identified several practical barriers. Session length was the lowest-rated attitudinal item across cohorts and the most variable, with respondents requesting a shorter session, longer mid-session break, or condensed introductory content. Respondents expressed a preference for in-person delivery to facilitate greater interactivity. Technical issues, including video playback quality and survey functionality, were noted by a few respondents. Gains were nonetheless consistent across all three cohorts. The feedback motivates in-person delivery, condensed pacing, and the integration of small group or case-based discussion in future iterations.

### Strengths and limitations

The principal strength is the consistency of the direction of effect across three iterations at a single site. This consistency mitigates some of the threats to internal validity inherent in a single-group pre-post design. The mixed-methods approach, combining quantitative pre-post measures with qualitative feedback, allows triangulation of findings and gives richer insight into the mechanisms driving the observed effects.

Several limitations should be acknowledged. First, 72% of Vevox participants (174 of 242) did not complete both surveys. As unsubmitted responses were not captured, non-completion reflects an unknown mixture of non-participation and mid-session attrition, and the potential for self-selection could not be assessed directly. Because participation was voluntary, students who completed both surveys may have been more engaged or receptive to the teaching than those who did not, potentially inflating the observed effects. Second, the single-group design cannot distinguish the session’s specific effects from those common to any structured teaching, such as expectation effects and demand characteristics. The immediate timing of the post-session survey means that gains may partly reflect short-term engagement with the topic rather than durable learning. Additionally, although students were instructed to complete the pre-session items before teaching began, the survey was accessible as a single form, making it possible for some participants to have completed all items after the session. This may have introduced response-shift bias^35^, potentially deflating pre-session ratings. Third, the outcome measures rely on self-reported confidence, which corresponds to Kirkpatrick’s Level 2 (learning) at most and does not capture whether participants subsequently changed their clinical behaviour (Level 3) or improved patient outcomes (Level 4)^36^. Furthermore, self-reported confidence may overestimate actual competence^37^. Fourth, the sample was drawn from a single institution, and the session was delivered online, so the findings may not generalise to other medical schools, or to in-person delivery where engagement dynamics differ. Fifth, the confidence questions were developed for this study and have not been independently validated. Finally, the intervention was designed, delivered, and evaluated by the same organisation, and two authors delivered the sessions. The analysis was undertaken by an author who took no part in delivery to reduce allegiance bias. Nevertheless, independent evaluation is needed to test whether these findings replicate.

### Implications for practice and research

The inclusion of gambling disorder in the MLA content map provides a policy lever for embedding gambling harm teaching. However, the uniformly low baseline confidence observed here suggests that inclusion in a content map alone is insufficient and structured teaching is needed to equip future clinicians to identify gambling harm in clinical practice. Although the present session was timetabled within the psychiatry rotation, gambling harm presents across general practice, emergency care, and safeguarding, as well as mental health contexts, and teaching need not be confined to a single specialty block.

Lived experience testimony was the most strongly endorsed element of the session in all three cohorts. The present design cannot isolate its contribution, as no comparison was made against equivalent teaching delivered without lived experience input, but the consistency of student endorsement suggests it merits consideration.

Several directions for future research emerge from this work. First, longitudinal follow-up during and beyond medical school is needed to determine whether confidence gains are sustained and whether gambling becomes a routine part of social history taking, as is the case with alcohol and smoking. Outcomes such as recording of gambling in clinical records and referral to support services are measurable only once participants enter practice and would require follow-up into the foundation years. Second, because medical curricula vary in structure and content, multi-institutional evaluation would test generalisability and identify how gambling harm teaching can best be incorporated within different curricular contexts. Third, comparative evaluations of different educational approaches are needed to establish which are most effective for teaching gambling harm. These may include experiential and simulation-based methods such as case-based discussion and simulated consultations, alongside the lived experience testimony and didactic teaching evaluated here.

## Conclusion

Across three cohorts, a single session delivered by individuals with lived experience and public health expertise was associated with significant gains in medical students’ self-reported confidence in recognising and responding to gambling harm. To our knowledge, this study is the first formal evaluation of gambling harm teaching for medical students. Whether these gains translate into changed clinical behaviour requires longer-term, multi-site evaluation with behavioural outcome measures. Given the scale of gambling harm and its systematic under-identification in clinical practice, medical schools should consider how gambling harm teaching might fit within their curricula and evaluate its introduction. The aim would be to normalise asking about gambling in routine social history taking, alongside alcohol and smoking.

## Competing interests

All authors are employed by Gambling Harm UK (Registered Charity Number 1196538), a national gambling harm prevention charity led by individuals with lived experience, which designed and delivered the intervention evaluated here. No author has received personal payment from any gambling operator, from GambleAware, or from any industry-affiliated body, and no author holds shares, consultancy arrangements, or any other financial interest in any gambling company.

Kishan Patel and Ben Jones delivered the sessions. Marriam Albukai conducted the analysis and drafted the manuscript but took no part in delivery.

Marriam Albukai is Director of Medical Education at Gambling Harm UK, by which she has been employed since November 2025. She is a medical student at King’s College London. She was on an interruption of studies throughout the period in which the sessions were delivered, was not a member of any participating cohort, and had no access to identifiable participant information.

Ben Jones is Director of Operations at Gambling Harm UK, by which he has been employed since May 2025.

Andrew Lovell is Medical Education Coordinator at Gambling Harm UK, by which he has been employed since December 2023.

Kishan Patel is a lived-experience co-founder of Gambling Harm UK and has been its CEO since July 2020. He also holds an NHS post as a Public Health Specialty Registrar (Thames Valley Deanery) and has undertaken gambling harm advocacy and policy activity since 2020, including written evidence to Parliament and responses to APPG and GMC consultations.

## Data statement

All data produced in the present work are contained in the manuscript and supplementary materials. Individual-level responses are not available, as participants were informed that access to their responses would be restricted to the research team.

