## Supplementary File 2 for "Evaluating the impact of a gambling harm focused educational intervention on medical students’ self-reported confidence: a non-randomised, single-group pre-post study"

### Pre-session

**1. I confirm that I have read and understood the Participant Information Sheet for the above study and have had the opportunity to ask questions.**

**Select a choice.**
Yes
No

### Pre-session

**2. I understand that my participation is voluntary and that I am free to withdraw at any time without giving a reason, before submitting my survey responses.**

**Select a choice.**
Yes
No

### Pre-session

**3. I understand that the study involves completing pre and post-teaching surveys to assess my knowledge and perceptions of gambling harm.**

**Select a choice.**
Yes
No

### Pre-session

**4. I understand that any information I provide will be anonymised and treated with strict confidentiality.**

**Select a choice.**
Yes
No

### Pre-session

**5. I understand that the anonymised survey responses may be used in academic reports, presentations, or publications.**

**Select a choice.**
Yes
No

### Pre-session

**6. I agree to take part in the study.**

**Select a choice.**
Yes
No

### Pre-session

#### 7. How confident are you that you can recognise when a patient might be experiencing gambling-related harm?

**Select a choice.**
Not at all confident
Slightly confident
Somewhat confident
Quite confident
Extremely confident

### Pre-session

#### 8. How confident are you that you can explain how gambling harm can affect a patient's health?

**Select a choice.**
Not at all confident
Slightly confident
Somewhat confident
Quite confident
Extremely confident

### Pre-session

#### 9. How confident are you that you can ask a patient about gambling in a sensitive and non-judgemental way?

**Select a choice.**
Not at all confident
Slightly confident
Somewhat confident
Quite confident
Extremely confident

### Pre-session

#### 10. How confident are you that you can identify appropriate referral or support options for a patient affected by gambling harm?

**Select a choice.**
Not at all confident
Slightly confident
Somewhat confident
Quite confident
Extremely confident

### Pre-session

#### 11. How confident are you that you can apply what you have learned about gambling harm in your clinical interactions with patients?

**Select a choice.**
Not at all confident
Slightly confident

Somewhat confident
Quite confident
Extremely confident

#### Pre-session

#### 12. How confident are you that you can consider gambling harm in population-level discussions?

**Select a choice.**
Not at all confident
Slightly confident
Somewhat confident
Quite confident
Extremely confident

#### Post-session

#### 13. How confident are you that you can recognise when a patient might be experiencing gambling-related harm?

**Select a choice.**
Not at all confident
Slightly confident
Somewhat confident
Quite confident
Extremely confident

#### Post-session

#### 14. How confident are you that you can explain how gambling harm can affect a patient's health?

**Select a choice.**
Not at all confident
Slightly confident
Somewhat confident
Quite confident
Extremely confident

#### Post-session

#### 15. How confident are you that you can ask a patient about gambling in a sensitive and non-judgemental way?

**Select a choice.**
Not at all confident
Slightly confident
Somewhat confident
Quite confident
Extremely confident

### Post-session

#### 16. How confident are you that you can identify appropriate referral or support options for a patient affected by gambling harm?

**Select a choice.**
Not at all confident
Slightly confident
Somewhat confident
Quite confident
Extremely confident

### Post-session

#### 17. How confident are you that you can apply what you have learned about gambling harm in your clinical interactions with patients?

**Select a choice.**
Not at all confident
Slightly confident
Somewhat confident
Quite confident
Extremely confident

### Post-session

#### 18. How confident are you that you can consider gambling harm in population-level discussions?

**Select a choice.**
Not at all confident
Slightly confident
Somewhat confident
Quite confident
Extremely confident

### Post-session

#### 19. Healthcare professionals have an important role in recognising and responding to gambling-related harm.

**Select a choice.**
Not at all confident
Slightly confident
Somewhat confident
Quite confident
Extremely confident

### Post-session

#### 20. Gambling-related harm is an important modifiable risk factor for health and wellbeing.

**Select a choice.**

Not at all confident
Slightly confident
Somewhat confident
Quite confident
Extremely confident

### Post-session

#### 21. Gambling-related harm is a neglected public-health concern.

**Select a choice.**
Not at all confident
Slightly confident
Somewhat confident
Quite confident
Extremely confident

### Post-session

#### 22. The session length was appropriate.

**Select a choice.**
Not at all confident
Slightly confident
Somewhat confident
Quite confident
Extremely confident

### Post-session

#### 23. The teaching materials were engaging and relevant.

**Select a choice.**
Not at all confident
Slightly confident
Somewhat confident
Quite confident
Extremely confident

### Post-session

#### 24. The session encouraged interaction and reflection.

**Select a choice.**
Not at all confident
Slightly confident
Somewhat confident
Quite confident
Extremely confident

### Post-session

#### 25. Hearing from individuals with lived experience enhanced my understanding of gambling harm.

**Select a choice.**
Not at all confident
Slightly confident
Somewhat confident
Quite confident
Extremely confident

### Post-session

### 26. I intend to apply what I learned in my work with patients.

**Select a choice.**
Not at all confident
Slightly confident
Somewhat confident
Quite confident
Extremely confident

### Post-session

### 27. I would recommend this teaching to other medical students.

**Select a choice.**
Not at all confident
Slightly confident
Somewhat confident
Quite confident
Extremely confident

### Post-session

### 28. What was the most valuable aspect of this session for you?

### Post-session

### 29. How could the session be improved for future students?
