## Supplementary Table 1 for "Evaluating the impact of a gambling harm focused educational intervention on medical students’ self-reported confidence: a non-randomised, single-group pre-post study"

**Supplementary Table 1.** Post-session attitudinal responses across three cohorts.

| Domain | Question | Cohort | n | Post-session Median Agreement (IQR) | % Agreement |
|---|---|---|---|---|---|
| Perceived importance of gambling harm | Healthcare professionals have a role in addressing gambling harm | Cohort 1 | 24 | 5.0 (4.8–5.0) | 100% |
| | | Cohort 2 | 20 | 5.0 (4.0–5.0) | 100% |
| | | Cohort 3 | 24 | 5.0 (5.0–5.0) | 100% |
| | Gambling harm is a modifiable risk factor | Cohort 1 | 24 | 5.0 (4.8–5.0) | 96% |
| | | Cohort 2 | 20 | 5.0 (4.8–5.0) | 100% |
| | | Cohort 3 | 24 | 5.0 (5.0–5.0) | 100% |
| | Gambling harm is a neglected public health concern | Cohort 1 | 24 | 5.0 (4.0–5.0) | 100% |
| | | Cohort 2 | 20 | 5.0 (4.0–5.0) | 100% |
| | | Cohort 3 | 24 | 5.0 (5.0–5.0) | 100% |
| Session quality and format | Session length was appropriate | Cohort 1 | 23 | 4.0 (4.0–4.0) | 91% |
| | | Cohort 2 | 20 | 4.0 (4.0–5.0) | 80% |
| | | Cohort 3 | 24 | 4.0 (4.0–5.0) | 100% |
| | Materials were engaging and relevant | Cohort 1 | 24 | 5.0 (4.0–5.0) | 92% |
| | | Cohort 2 | 20 | 5.0 (4.0–5.0) | 100% |
| | | Cohort 3 | 23 | 5.0 (4.0–5.0) | 100% |
| | Session encouraged interaction | Cohort 1 | 24 | 5.0 (4.0–5.0) | 96% |
| | | Cohort 2 | 19 | 5.0 (4.0–5.0) | 95% |
| | | Cohort 3 | 24 | 5.0 (4.0–5.0) | 100% |
| Value of lived experience | Lived experience enhanced my understanding | Cohort 1 | 24 | 5.0 (5.0–5.0) | 100% |
| | | Cohort 2 | 20 | 5.0 (5.0–5.0) | 95% |
| | | Cohort 3 | 24 | 5.0 (5.0–5.0) | 100% |
| Intention and endorsement of the session | I intend to apply this learning clinically | Cohort 1 | 24 | 5.0 (4.8–5.0) | 100% |
| | | Cohort 2 | 20 | 5.0 (4.0–5.0) | 100% |
| | | Cohort 3 | 23 | 5.0 (5.0–5.0) | 100% |
| | I would recommend this session to peers | Cohort 1 | 24 | 5.0 (4.0–5.0) | 100% |
| | | Cohort 2 | 20 | 5.0 (4.0–5.0) | 100% |
| | | Cohort 3 | 24 | 5.0 (5.0–5.0) | 100% |

Responses were rated on a five-point Likert scale: 1 = Strongly disagree, 2 = Disagree, 3 = Neither agree nor disagree, 4 = Agree, 5 = Strongly agree.

% Agreement reflects the proportion of respondents selecting “Agree” or “Strongly agree”.

n = number of responses; IQR = interquartile range
